# A global analysis of quarantine and isolation policies governing outbreak responses

**DOI:** 10.1101/2024.11.20.24317642

**Authors:** Amanda Rosner, Ciara M. Weets, Rory Wilson, Rebecca Katz

**Affiliations:** Georgetown University Center for Global Health Science and Security, 3900 Reservoir Road NW Washington, DC 20057

## Abstract

**Introduction:** Countries across the world implemented diverse quarantine and isolation policies throughout the COVID-19 pandemic with varying levels of effectiveness. Their widespread use invites new considerations regarding the effectiveness of domestic quarantine and isolation policies, the ways they are enforced, and the jurisdictions responsible for ordering these measures.

**Methods:** We systematically analyzed legally-enforceable policies in current standing in each United Nations (UN) member state, assessing the authorities to quarantine and isolate individuals within national borders. We captured the text of each policy and categorized the responsible jurisdictional authority and enforcement mechanisms.

**Results:** Of UN member states, 91.67% (176/192) had legally-enforceable policies that addressed both quarantine and isolation. Two countries only had quarantine policies, seven only had isolation policies, and seven countries had neither. Jurisdictional quarantine authority was primarily vested in the national level (74.16%; 132/178), with the remainder mixed (22.47%; 40/178) and subnational only (3.37%; 6/178). Isolation authority was also primarily at the national level (69.40%; 127/183) but with a greater proportion mixed (27.87%; 51/183) and subnational only (2.73%; 5/183).

Quarantine enforcement mechanisms were codified in a majority of countries (80.91%; 144/178) with nearly all (94.44%; 136/144) enforcing quarantine through monetary fines or incarceration penalties for non-compliant individuals. Isolation enforcement mechanisms were codified in an even greater number of countries (86.89%; 159/183), with 95.51% (149/156) having penalties for non-compliant individuals.

**Conclusion:** We created a novel repository for quarantine and isolation policies to assist in future outbreak responses. We identify specific country-level policy gaps, which can be addressed through epidemic and pandemic preparedness efforts. Finally, the repository provides the necessary evidence base for future research analyzing the impact of quarantine and isolation policies upon disease outbreak response outcomes.

**Key Messages:** *1. What is already known on this topic - summarize the state of scientific knowledge on this subject before you did your study and why this study needed to be done*

Non-pharmaceutical interventions, including quarantine and isolation, are critical to controlling the spread of infectious diseases. Legally-enforceable policies often authorize jurisdictional authorities to implement and enforce non-pharmaceutical intervention strategies. During the COVID-19 pandemic, there was widespread and diverse implementation of domestic quarantine and isolation policies. However, there have been no global efforts to collect and analyze the legal frameworks for quarantine and isolation governance and enforcement authorities.

*2. What this study adds - summarize what we now know as a result of this study that we did not know before*

We comprehensively map the current regulatory environment for legally-enforceable quarantine and isolation policies in each UN Member State. We use representative policy examples to demonstrate the diversity in jurisdictional authority to order quarantine and isolation, as well analyze the current penalty schemes employed by nations to enforce non-pharmaceutical interventions. We find that financial enforcement mechanisms that reflect economic fluctuations may remain a more durable deterrent overtime than set penalty ranges, yet few countries utilize such flexible penalization schemes.

*3. How this study might affect research, practice or policy - summarize the implications of this study*

Our novel repository of global quarantine and isolation policies can be used to assist future outbreak responses and identify country-level policy gaps to be addressed through preparedness efforts and updating legal frameworks. Analysis of the impact of different quarantine and isolation policies and enforcement mechanisms upon disease outbreak outcomes using this repository are warranted.

## Introduction

Quarantine and isolation policies are critically important public health measures to control infectious disease outbreaks. Quarantine–the separation of persons exposed to an infectious disease–and isolation–the separation of individuals who are symptomatic or are confirmed to have an infectious disease–have been used throughout history to manage outbreaks, with passages from the Old Testament suggesting that isolation of “unclean” individuals was commonplace [1]. The use of both quarantine and isolation and the codification of associated policies have been well documented over the millennia, from the Plague of Justinian to the Venetian practice of holding ships at anchor in the fourteenth century [2,3].

In the modern context, policies typically make national governments responsible for quarantine and isolation at their own international borders [4]. Countries typically have additional wording or separate policies granting quarantine and isolation authority within their national borders to applicable levels of government. Quarantine and isolation policies were widely invoked around the world during the COVID-19 pandemic [5]. However, the implementation of these policies by different countries during the pandemic was diverse and resulted in varying levels of effectiveness [6]. This widespread use of quarantine and isolation invites new considerations regarding the effectiveness of policies, the ways they are enforced, and the jurisdictions responsible for ordering these measures.

In domestic outbreak responses, having a clear understanding as to who has the authority to quarantine or isolate in different situations and how they can enforce these measures are critical for timely action. On an international level, understanding which policies provide quarantine and isolation authority can help coordinate global responses to outbreaks and help identify country-level policy gaps to assist epidemic and pandemic preparedness efforts.

We aimed to identify the domestic quarantine and isolation policies in each United Nations (UN) member state, the level of government authorized to carry out these measures, and what enforcement mechanisms are available. These efforts build the evidence base necessary to enable research into how quarantine and isolation policies impact outbreak response outcomes.

Additionally, this research will provide a repository for quarantine and isolation policies which can be used by national or international actors to assist future outbreak responses.

## Methods

This research was conducted as part of the Analysis and Mapping of Policies for Emerging Infectious Diseases project (AMP EID), for which subject matter experts systematically surface, collect, and assess legally-enforceable health-related policies from each UN member state [7].

We included all UN member states in this study, with the exception of the Democratic People’s Republic of North Korea; this country was omitted due to the research team’s inability to access and verify policy documents [8].

We followed a standardized operating procedure for data collection, beginning with a literature review, where the research team identified applicable search terms to use when surfacing policies for each country. Findings from a landscape review informed the development of research questions and the data taxonomy (Supplementary Materials). The research team then conducted a sample study on 10 countries, selected to represent diverse governmental systems, economic statuses, and geographic locations. After the completion of this sample study, the team resolved any gaps in the data collection protocol, data taxonomy, and coding methodology.

### Definitions

Throughout our research, we only looked at the authority to quarantine and isolate humans within national borders. We excluded quarantine and isolation provisions about animals, cargo, international borders, international travelers, and ships. For our research, *patient* refers to the sick person in instances of isolation, and *contact* refers to the person exposed to a disease for instances of quarantine. Our standardized definitions are available in Supplementary Materials (Supplementary Table 1).

### Policy Identification

We identified policies through a protocol that encompassed reviewing existing document repositories, standardized searches of peer-reviewed publication platforms, search engines, and digitized national legal databases.

We first inspected known repositories for health policies, including FAOLEX and geographic-specific policy repositories such as Droit Afrique and PACLII [9,10,11]. A standardized search using Google Scholar and the broader Google search engine was then run using the series of queries developed during the literature review and tested by the sample study in countries where the government operates and publishes laws in a language other than English; machine-translation was used to translate the search terms into the official language(s) of the country. For non-English-speaking nations, all searches were first run in English, and then all translated search terms were run, altering the word order within each term or substituting a synonym as needed, to make the search structure appropriate for the language into which it was translated.

When search engines failed to directly surface a policy, yet peer-reviewed sources yielded evidence of an applicable document, the identified document of interest was individually searched for either by title or through other search terms relevant only to that policy.

After reviewing available peer-reviewed and gray literature, in addition to documents included in previously compiled policy databases, the research team searched national governmental websites where policies might be located. The standardized search terms were employed using search functions on the websites (Supplementary Materials: Search Terms). In the case that there was no available search capacity on the website, the research team manually reviewed the repository for keywords related to the topic. If no potentially-relevant policies were discovered through these processes, the researchers consulted constitutional case law to see if authority was vested through vague language in the constitution but had been used to allow quarantine and/or isolation in practice. On several occasions, governments were contacted directly to inquire about the presence or the content of a policy.

### Screening and Creation of the Quarantine and Isolation Database

All policies were reviewed against inclusion criteria to ensure they had current standing and were legally enforceable (Supplementary Figures 1-3; Supplementary Table 1). Plans, strategies, or guidance documents were excluded. Likewise, laws and other policies created and implemented only for the COVID-19 pandemic, but that were no longer enforceable, were excluded from this dataset when there were other standing, legally-enforceable policies in place. In the cases where there were no identifiable standing policies for quarantine and/or isolation in a country, we included laws developed and enforced during the COVID-19 pandemic, as a demonstration of the legal course of action a country has previously taken during an infectious disease outbreak to implement either or both of these policies.

Included policies were downloaded as PDFs and captured in Airtable, a cloud-based relational database platform [12]. For each country, policies were qualitatively coded for each subtopic. All policies were collected between November 2023 and September 2024. Where necessary, documents were translated using machine translation. When possible, policy documents were interpreted and coded by researchers fluent in the given language, while fluent speakers were contacted to verify machine translations for languages not spoken by the research team.

### Data Validation

For quality assurance and control, the research team had a secondary reviewer use Perplexity AI, an artificial intelligence search engine, to assess the concordance of the results [13]. Any results that were non-concordant were reviewed by the research team and reconciled based upon the available information and the inclusion criteria.

### Data Availability Statement

The policy repository and country status data are available at https://github.com/cghss/Quar-Iso.

## Results

Across 192 UN member states, we found that 91.67% (176/192) of them had policies that addressed both quarantine and isolation. Two countries had policies for quarantine but not for isolation. Seven countries had policies for isolation but not quarantine. We found no legally-enforceable policies for either quarantine or isolation in seven countries, namely: Central African Republic, Eritrea, Mauritania, Republic of Congo, Rwanda, Somalia, and South Sudan [14]. The seven countries which lacked both quarantine and isolation policies were all in Africa. Three of these countries shared borders: the Republic of Congo, Central African Republic, and South Sudan. For 13 of the 192 countries, we captured policies developed in response to the COVID-19 pandemic as a proxy for standing legislation or if they replaced antiquated laws and/or supplemented existing health policies.

### Jurisdictional Authority for Quarantine and Isolation

Since different countries assign the authority to quarantine or isolate to different levels of government, we created three categories of governments that could have quarantine or isolation authority: national, subnational, and mixed. Authority to quarantine was identified in 92.71% countries (178/192). The vast majority of countries with quarantine authority (74.16%; 132/178) vested quarantine authority at the national level. A further 22.47% (40/178) of countries had mixed authority, maintaining some aspect of national quarantine and devolving some quarantine authority to subnational levels, like in the United States, where Article X of the Constitution grants quarantine and isolation authority to the states and Title 42 of the United States Code bestows authority to the federal government for specific diseases in scenarios that fall under federal jurisdiction [15,16,17]. The complete delegation of domestic quarantine authority to subnational jurisdictions was uncommon, occurring in only 3.37% (6/178) of countries. Within these six countries, there was notable variation in where the subnational power to quarantine rests: exclusively intermediate-level governments for four countries; solely local governments for only one country; and a combination of intermediate and local governments for only one country.

While the majority of countries had policies allowing for both government mandated quarantine and isolation, there was variation in the target of these interventions. Quarantine can be imposed on individuals or it can take place at a population-level through *cordon sanitaire*, which is a series of measures that can be implemented to prevent groups of people from exiting a geographic area. We found that 21 of the countries with quarantine authority (11.8%; 21/178) allowed for both individual and population-level quarantine, allowing the government to enforce quarantine orders on certain persons and/or geographically distributed sections of the population. In three countries, the only codified method of quarantine was at the population-level.

Isolation authority was found in 183 countries (95.31%; 183/192). In the majority of countries, isolation authority was found to be vested in only the national level government. There were 127 countries (69.40%; 127/183) with solely national-level authority for isolation; 51 countries (27.87%; 51/183) had isolation authority shared between national and subnational governments, depending on specific scenarios that were articulated in the respective policies. The remaining five countries, all of which also vested quarantine powers within subnational governing bodies, vested the power to isolate in subnational jurisdictions. Only 13 countries had different levels of government in charge of quarantine than they did for isolation.

### Enforcement Mechanisms for Quarantine and Isolation

Various mechanisms were found to enforce quarantine and isolation around the world, including penalties for the violation of quarantine or isolation orders, the use of force, arrest, and forced confinement. Enforcement mechanisms were codified in 80.91% (144/178) of countries with quarantine authority. Nearly all of these countries (94.44%; 136/144) enforced quarantine orders by imposing penalties for noncompliance on the quarantined individuals. A further 2.78% (4/144) of countries devolved the authority for enforcing quarantine to subnational governments, creating heterogeneity in enforcement mechanisms across subnational jurisdictions. The remaining countries included provisions for penalizing overseeing physicians when they fail to quarantine contacts of infectious diseases. In three countries, penalties for noncompliance were imposed on both the contact and the physician. In one country, Czechia, the law penalized physicians who fail to order quarantine but did not penalize contacts who violate quarantine orders. Notably, 34 countries with quarantine authority did not have quarantine enforcement mechanisms.

The majority of countries (64%; 123/192) utilized a monetary fine and/or imprisonment to enforce quarantine. Seventeen of these countries state that one of these penalties would be enforced for non-compliance, but they did not define the value of the penalty in existing policy. A further six countries utilized monetary penalties specific to an individual’s daily wage or base salary. Such is the case in Costa Rica, which penalizes individuals for disregarding quarantine orders through a fine of three base salaries [18]. Similarly, six countries dictate a number of penalty units that may be assigned to individuals that break quarantine orders. Ghanian policy, for instance, orders that individuals that engage such indiscretions be assigned a fine of up to 50 penalty units [19]. A total of six countries listed their monetary penalties in a currency that is no longer used in the country. For example, the financial penalty for breaking quarantine or isolation orders in Italy is listed in Lira, a unit of currency that has not exclusively been used in the country since 1999 [20].

For countries that specify a monetary fine or incarceration period for violating quarantine orders, we found substantial variation in the penalties across different nations. South Africa and Namibia allow individuals to be sentenced to up to 10 years in jail for breaking quarantine, while Australia and the Netherlands can impose fines greater than $90,000.00 USD. These penalties are outliers, as the median value for a fine was $200.14 USD. In 25 countries, the maximum penalty was less than $100.00 USD, while 11 of these nations imposed a maximum fine of less than $5.00 USD. The maximum fine values ranged from less than one cent USD and nearly $100,000.00 USD. Likewise, the median jail sentence was less than one month. Maximum values for length of imprisonment ranged from 15 days to 10 years. Additionally, some countries have enforcement mechanisms besides monetary fines and/or imprisonment, either in addition to fine/imprisonment or in place thereof. Examples of these other enforcement mechanisms included police involvement, arrest, and forcibly quarantining the contact.

Enforcement mechanisms for isolation were present in a higher proportion of countries than quarantine enforcement mechanisms. Of the 183 countries with legally-enforceable isolation policies, 159 (86.89%) had a codified enforcement mechanism, while the remaining 24 did not include such mechanisms in policy. Three countries devolved isolation enforcement powers to subnational jurisdictions, for which each jurisdiction’s enforcement mechanisms can differ. Within the remaining 156 nations that enforced isolation in national level policy, the vast majority (95.51%; 149/156) imposed penalties solely on noncompliant individuals placed under isolation. Broader isolation enforcement mechanisms for both patients and physicians were in place in six countries, all of which are in Europe and half of which also have quarantine enforcement mechanisms for both patients and physicians.

We found that 131 countries penalize violations of isolation orders through monetary fine and/or incarceration. There were 20 countries with a different penalty for breaking isolation than for breaking quarantine requirements. The median value of the fine for breaking isolation was $124.10 USD, with penalties ranging from less than one tenth of a cent USD, and $99,000.00 USD. Thirty-five countries had a fine less than $100.00 USD, with 20 of these countries including a fine in current policy that was valued at less than $12.00 USD. There were an average of nine months of incarceration permitted in legislation, with maximum sentences ranging from 15 days to 10 years.

## Discussion

### Jurisdictional Authority

Across the world, we found substantial variation in which jurisdictions had the authority to quarantine and isolate, as well as which enforcement mechanisms could be used to ensure compliance. Notably, we also discovered 23 countries without quarantine and/or isolation authority. While this research captures the policies that are codified, it does not capture how the policies are implemented, or where some jurisdictions may have taken action during a public health emergency in the absence of a specific, legally enforceable policy.

The COVID-19 pandemic has provided an opportunity to study the effectiveness of quarantine and isolation in a variety of scenarios. This is particularly relevant to which jurisdictional authorities are the most effective, given there have been growing calls for increased localization within public health emergency responses [21,22,23]. However, we found only 46 subnational governments had any authority for quarantine and only 56 had any authority for isolation, combining both subnational and mixed structures. Our repository of policies will now allow for analyses to determine if subnational jurisdiction for quarantine and isolation led to better outcomes during public health emergencies, examining morbidity, mortality, protection of vulnerable populations, and population resilience.

### Enforcement Mechanisms

Enforcement mechanisms for quarantine and isolation are a critical balance between ensuring the population follows public health instructions, maintaining public order, ensuring trust in government, and refraining from criminalizing illness. At their core, enforcement mechanisms influence the culture of quarantine and isolation, as well as the compact between individuals and their government [24].

Our findings on enforcement mechanisms for quarantine and isolation were notable in part due to their large variation. The main two forms of enforcement mechanisms for both quarantine and isolation were financial penalties and/or imprisonment for the contact and patient, respectively, who violated their quarantine or isolation order. There were occasionally physical enforcement mechanisms besides financial penalties and imprisonment, including the use of force and locking doors from the outside. There was a mix between these physical barriers as standalone enforcement mechanisms and when they were paired with fines and/or imprisonment. Additionally, some countries penalize providers for not adhering to isolation policies.

Determining which enforcement mechanisms are the most effective requires determining how the strength of enforcement mechanisms differ across countries. A standard financial penalty across countries will carry different weights due to differences in average household incomes, cost of living, and other economic measures across countries. For example, some nations have adopted enforcement policies based on fee schedules or penalty units that change over time. A fine equal to a daily wage will be variable in amount, but may have the same intended impact on the individual over decades. Other countries have put a set fine in place, as determined as an appropriate incentive for action at a single point in time. In many cases, these fines have not aged well, having not kept pace with inflation.

It is also unclear if equal time periods of imprisonment carry the same weight as a disincentive across countries and penal systems. Another important consideration is how the triggers for enacting enforcement mechanisms differ between countries. As scholarship is developed to identify the levels of enforcement, fines and punishment that incentivize behavior in different populations, special consideration should be given to how enforcement mechanisms are articulated in policies and laws to have the intended impact over time.

## Limitations

The methodology employed in this research has several limitations. When reviewing policies in languages not spoken by the research team, we relied on translation software, which may be inaccurate at times. Precise translation is critical, as to accurately interpret the legal meaning of each word, punctuation, and the grammatical structure. Additionally, we were limited to the literal interpretation of these policies, especially for the often-misused terms of *quarantine* and *isolation*. The terms *quarantine* and *isolation* are frequently conflated, particularly in the policies enacted during and after the COVID-19 pandemic [25]. There were also instances where laws were open to interpretation regarding the definitions of quarantine and isolation [26,27].

At the same time, non-specific language left many policies open for interpretation. Because of this, we documented the rationale behind our decisions in the dataset as status justifications, and include the text of the policy in our dataset so others may apply different approaches to interpretation. Finally, we note the limitation that our research only captures one point in time and the policies might change in the future.

## Conclusion

Our research creates the necessary evidence base to enable investigation into the impact of quarantine and isolation policies on disease outbreak response outcomes. It also serves as a repository for quarantine and isolation policies to assist in future outbreak responses. Our results showed country-level policy gaps where there are no enforcement mechanisms, no quarantine authority, or no isolation authority. These gaps can be addressed by domestic and international actors as part of preparedness efforts. We also revealed future avenues where further research on quarantine and isolation policies is critical.

## Data Availability

Data are available in a public, open access repository.

https://github.com/cghss/Quar-Iso

## Acknowledgments

This research was funded by a grant from Rockefeller Foundation. The funder had no part in the conceptualization, design or analysis of the study. The authors would like to acknowledge the assistance of Zahra Izzi in creating the supplemental figures. We would also like to acknowledge Ellie Graeden, Tess Stevens, Hailey Robertson, and Ryan Zimmerman in contributing to designing of the data structure, and managing of the project database.

---

**Figure 1.**
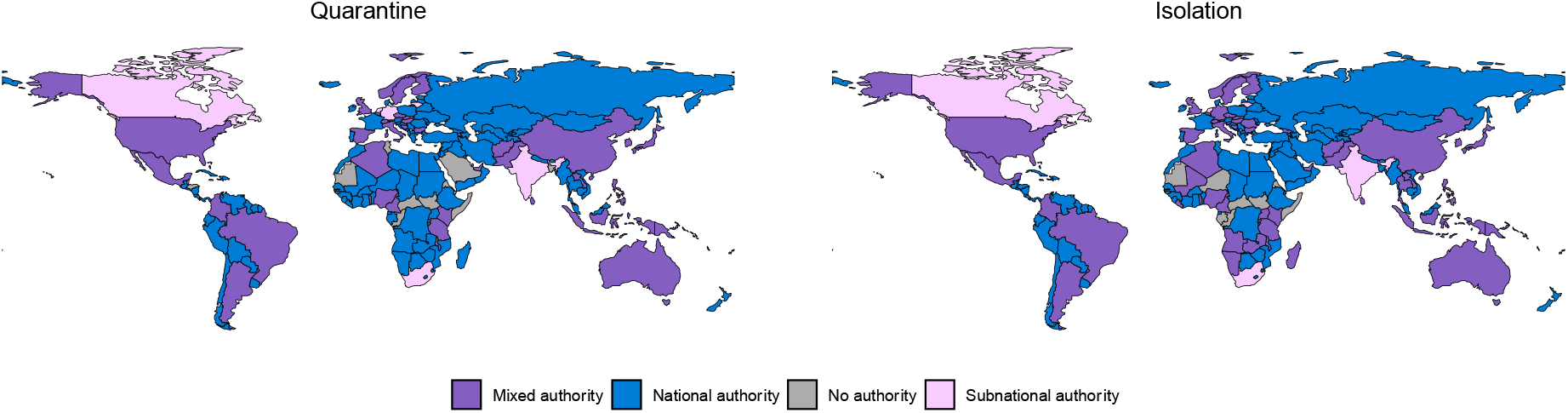
Global Mapping of Countries with Quarantine and Isolation Authorities. Jurisdictional authority for quarantine and isolation were determined by analysis of legally-enforceable policies. Mixed authority refers to instances in which the authority to implement and enforce non-pharmaceutical interventions were vested in national and subnational governing bodies.

**Figure 2.**
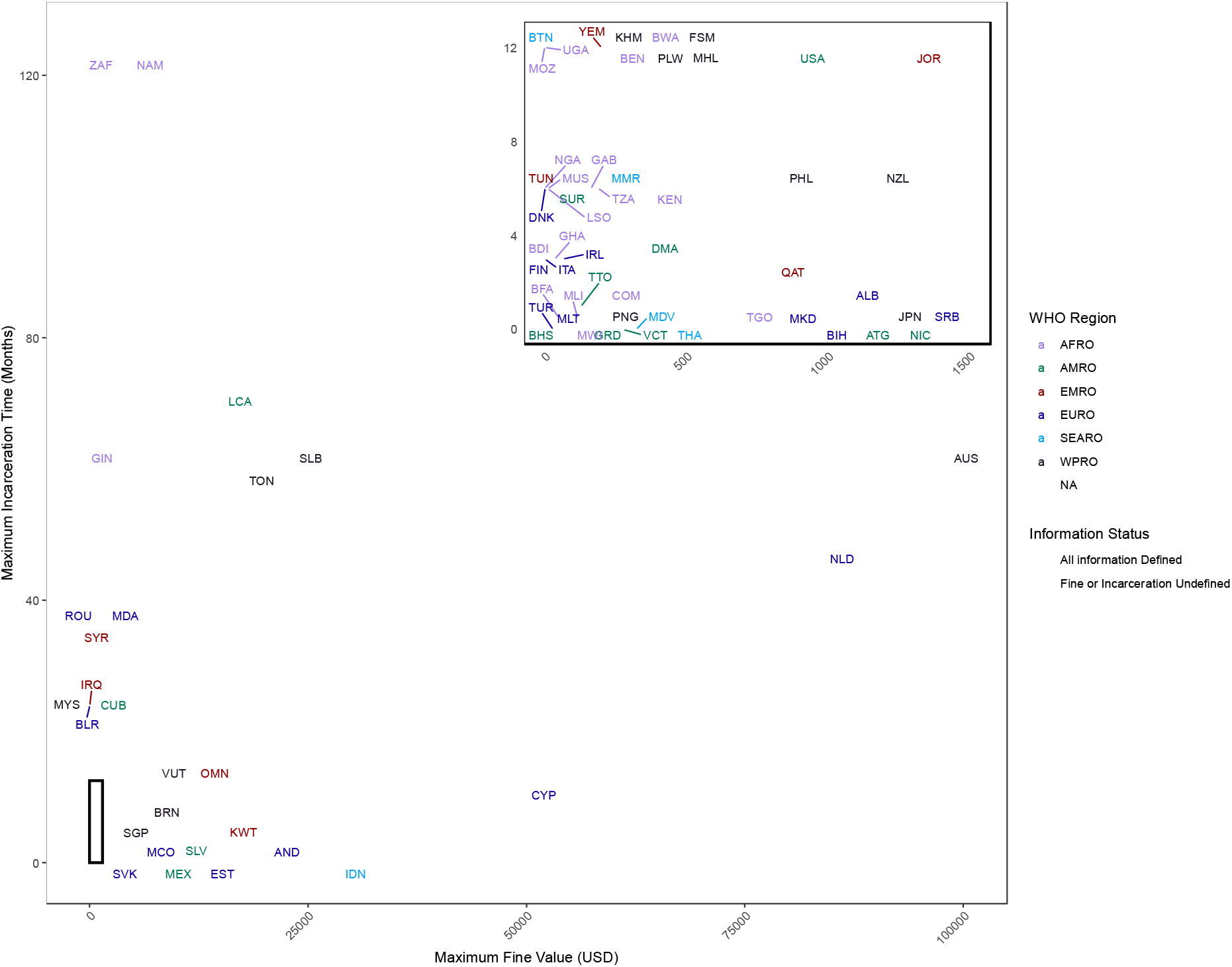
National-Level Quarantine Enforcement Mechanisms: Fines and Incarceration. Penalty data were extracted from relevant legally-enforceable policies. Monetary fines were converted from domestic currency to US Dollars using the exchange rate on November 7, 2024. Countries are denoted by the International Standardization Organization (ISO) 3166 country codes; colors were assigned based upon World Health Organization regions. Nations designated by a triangle include the opportunity for both types of penalty to be invoked, yet leave one of the two values unspecified. All unspecified entries were assigned the value 1.

**Figure 3.**
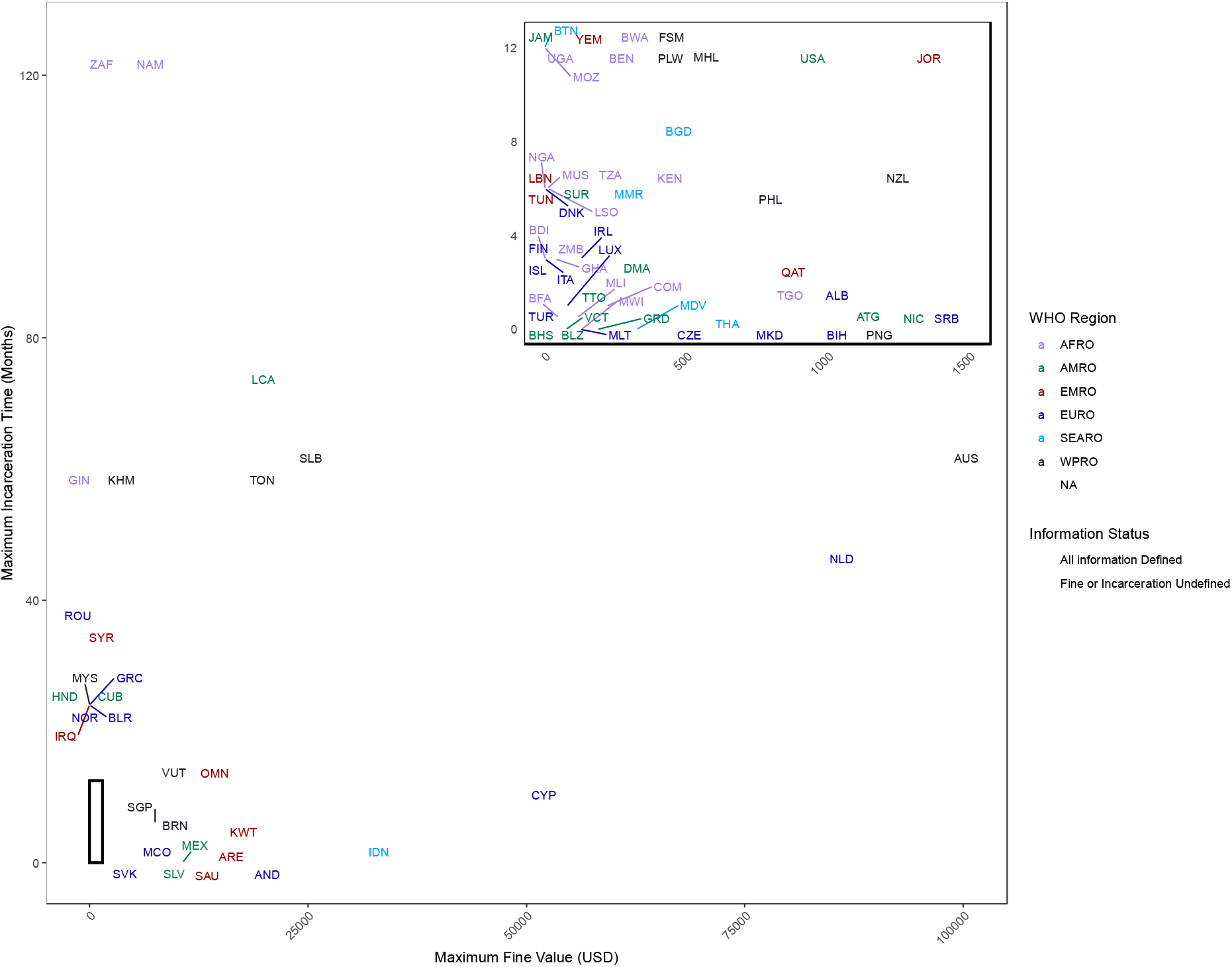
National-Level Isolation Enforcement Mechanisms: Fines and Incarceration. Penalty data were extracted from relevant legally-enforceable policies. Monetary fines were converted from domestic currency to US Dollars using the exchange rate on November 7, 2024. Countries are denoted by the International Standardization Organization (ISO) 3166 country codes; colors were assigned based upon World Health Organization regions. Nations designated by a triangle include the opportunity for both types of penalty to be invoked, yet leave one of the two values unspecified. All unspecified entries were assigned the value 1.

