## Supplementary Figures 1-3 for "A global analysis of quarantine and isolation policies governing outbreak responses"

**Supplementary Materials**

**Supplementary Figure 1: Quarantine and Isolation Policy Status**

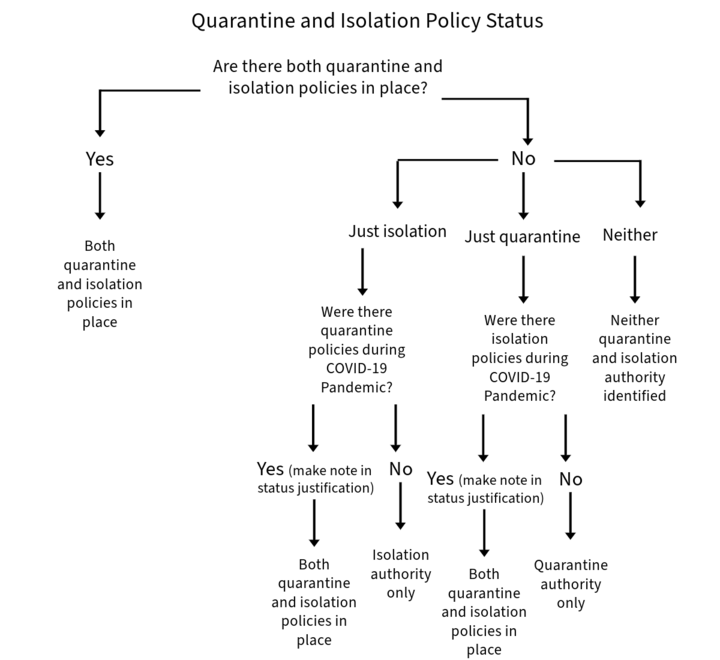

**Supplementary Figure 2: Quarantine/Isolation Authority Level**
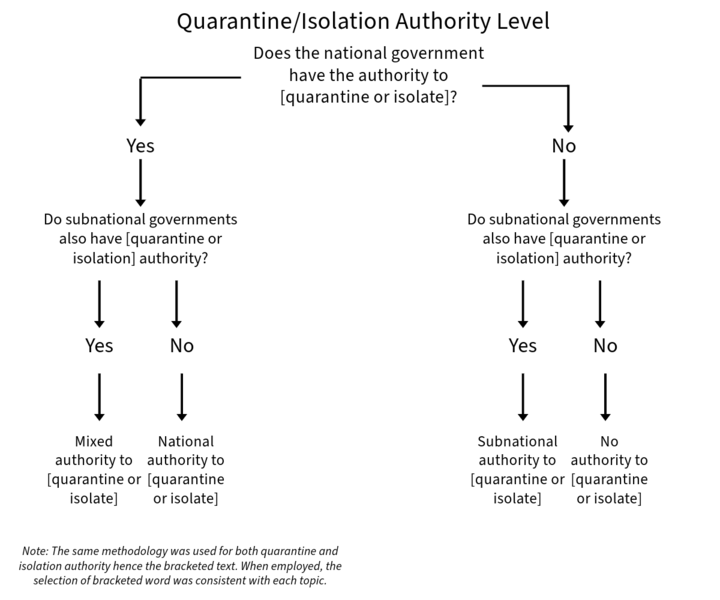

**Supplementary Figure 3: Coding Methodology for Quarantine/Isolation Enforcement Mechanisms**
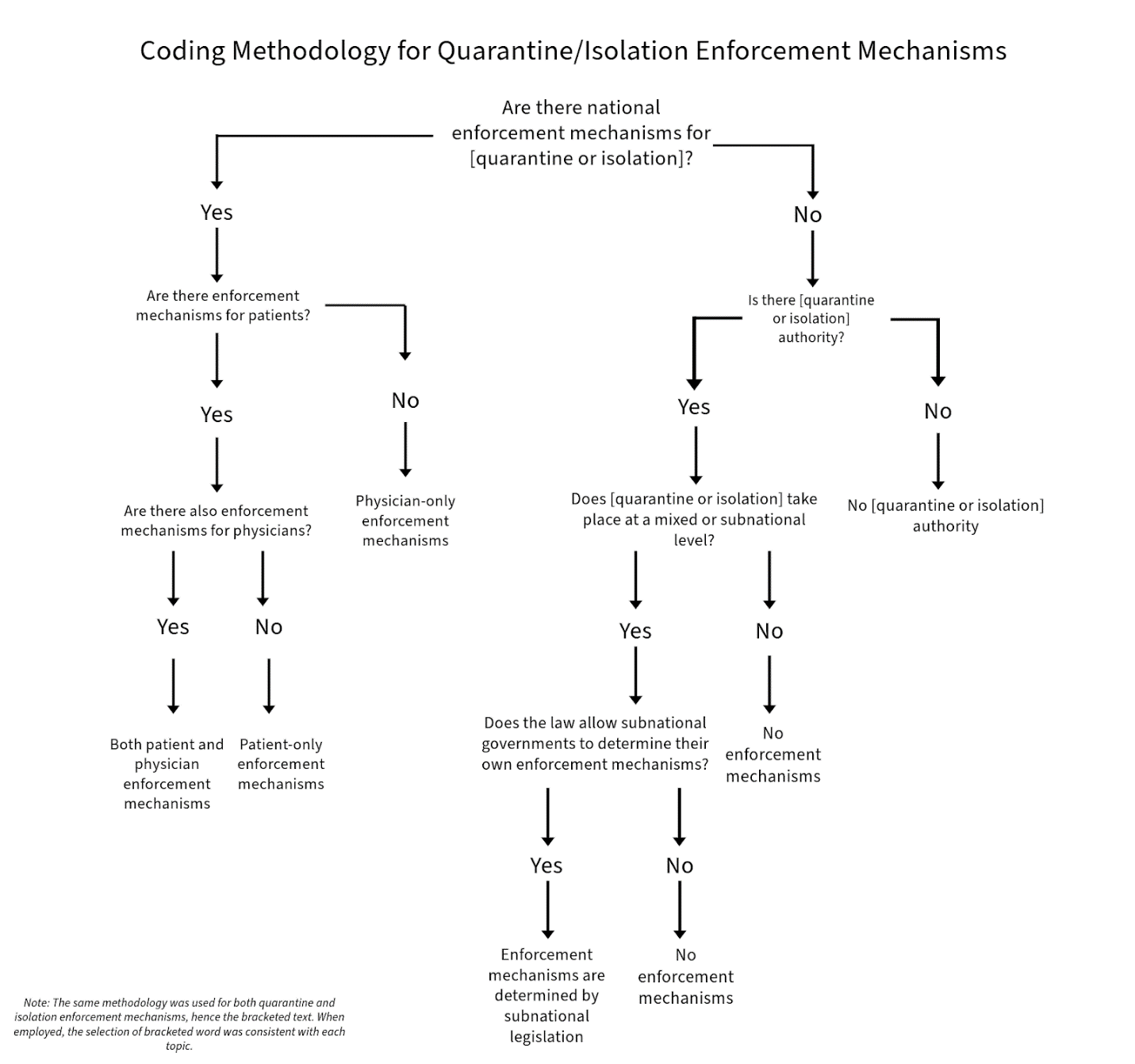

**Supplementary Table 1. Definitions and Policy Inclusion Criteria**

| **Term** | **Definition and Criteria** |
| --- | --- |
| Contact | A person who has been in contact with or exposed to someone with an infectious disease. |
| Enforcement Mechanisms | We define *enforcement mechanisms* to include measures used to enforce quarantine or isolation, including measures intended to physically force someone to quarantine/isolate, punishment of quarantine/isolation violation, and/or measures to ensure that physicians quarantine/isolate their contact or patient.  Contact/patient enforcement mechanisms include penalties for violating quarantine/isolation orders, respectively, such as fines, imprisonment, or forced confinement. Additionally, there can be physical enforcement mechanisms to ensure compliance, such as forced confinement, use of force, locking doors from outside, and returning someone to a site of quarantine/isolation. Notably, forced medical treatment does not count as a patient-only enforcement mechanism, unless the forced medical treatment only applies to persons who have violated orders.  Physician enforcement mechanisms include penalties for the failure of a physician to quarantine/isolate a contact or patient. These can include financial penalties, imprisonment, loss of license, or etc. We did not count penalties for a physician’s failure to report a disease to authorities as physician-only quarantine/isolation enforcement mechanisms. |
| Intermediate Governments | *Intermediate government* refers to a level of government between national and local government, such as states, provinces, or other equivalents. |
| Isolation | *Isolation* refers to the series of measures to separate an ill person from other individuals. Under our definition of *isolation*, these measures apply to ill persons, symptomatic persons who are suspected of being but not yet confirmed to be sick, and asymptomatic patients who are confirmed to be infected or are carriers.  Unless proof was provided of the government using a different interpretation, we classified descriptions of forced hospitalization as isolation, since hospitalization typically implies a sick patient. Additionally, isolation could take place in a hospital, specialized facility, or the patient’s home.  We included instances of broad governmental authority to take measures in the field of disease prevention as the authority to isolate, as well as the authority to take prophylaxis measures when not otherwise specified.  Finally, our definition of *isolation* excludes all isolation situations relating to international borders, travelers, and maritime quarantine. |
| Local Government | *Local governments* encompass municipal, village, town, county, and equivalent levels of government. |
| Mixed Authority | Governments were classified as having *mixed authority* to quarantine/isolate when both subnational and national governments had the authority to quarantine/isolate, whether it be across the same conditions or under different circumstances. |
| National Government | The term *national government* included any of the following representatives or their equivalents: national government ministries; government minister; kings; government physicians employed by the national government; physicians who face penalties from the national government for the failure to quarantine/isolate a contact or patient; national courts; persons appointed by or authorized by the national government.  National authorities also encompassed instances where no specific authorities were named and when broad terms like “health authorities” were used and not otherwise specified. Additionally, national authorities encompass instances when country governments were given broad authority and responsibilities for disease control measures. |
| Patient | *Patient* refers to the sick person in instances of isolation. |
| Physician | *Physician* encompasses whichever level of medical providers determined in law (whether that be physician, or physician-equivalent) who are involved in the quarantine and isolation process. |
| Physician Authority | Physician authority to quarantine/isolate is when medical providers have the authority to quarantine/isolate, either standalone or in combination with the government. Physician authority to quarantine/isolate also encompasses the occasions where physicians are given temporary authority to quarantine/isolate, while the government initiates the formal process to quarantine/isolate.  Physician authority to quarantine/isolate was noted but not treated as a standalone category.  Physician authority does not include the authority of government-employed physicians to quarantine/isolate; there, the authority is vested in whatever level of government employs the physician. |
| Policy | For our research, *policies* refer to legally-enforceable documents, orders, decrees, regulations, and etc.  For this paper, policies refer to standing and current documents, except when standing policies were unavailable for quarantine/isolation. In those instances, COVID-19 policies were used in place of standing policies, when available. The only COVID-19 policies that counted as standing policies are policies that are either still on the books, include COVID-19 as one of multiple diseases for which the law is about, and/or any permanent amendments to other policies that added COVID-19-specific provisions. |
| Quarantine | *Quarantine* refers to the series of measures to separate someone who has been exposed to an illness, but is not (or not yet) sick, from other individuals. Under our definition of *quarantine*, these measures apply to contacts of infectious disease patients and persons who are presumed to have been exposed to an infectious disease.  Unless proof was provided of the government forced hospitalization as a form of quarantine, we did not count forced hospitalization as quarantine, since hospitalization typically implies a sick patient, and therefore falls under isolation rather than quarantine. Additionally, our definition allowed for quarantine that could take place at a specialized facility or at a contact’s home.  We included instances of broad governmental authority to take measures in the field of disease prevention as the authority to quarantine, as well as the authority to take prophylaxis measures when not otherwise specified.  Our definition of *quarantine* measures counted being held for medical observation, but it did not include forced medical exams, as a forced medical examination does not keep the contact separate from the rest of the population for a prolonged period of time.  Additionally, our definition of quarantine included both the quarantine of individuals and the institution of population-level quarantine measures, or *cordon sanitaire*. Population-level quarantine is the series of measures that can be implemented to prevent people from exiting an “infected area.”  Finally, our definition of *quarantine* excludes all quarantine situations relating to international borders, travelers, and maritime quarantine. |
| Subnational Government | *Subnational governments* refer to any level of government that falls under the national government, such as local governments and/or intermediate governments, when applicable.  Subnational quarantine/isolation authority includes when states, provinces, regions, subnational courts, subnational government officials, persons appointed by subnational governments, and/or any combination of subnational-level authorities have the power to quarantine/isolate under at least one set of circumstances. Subnational government officials or persons appointed by subnational governments. |

**Supplementary Material: Search Terms**

Quarantine authority law, quarantine law, isolation authority law, isolation law, isolation policy, quarantine policy, disease isolation authority law, disease isolation law, disease isolation policy, infectious disease law, infectious disease policy, disease law, public health law, public health code, public health policy, sanitation code, hygiene code, health law, epidemic law, epidemic policy, tuberculosis law, tuberculosis isolation law, TB isolation law, TB law, leprosy law, leprosy isolation law, leper law, Hansen’s disease Law, Hansen disease law.
